# Validation and comparison of PICTURE analytic and Epic Deterioration Index for COVID-19

**DOI:** 10.1101/2020.07.08.20145078

**Authors:** Brandon C. Cummings, Sardar Ansari, Jonathan R. Motyka, Guan Wang, Richard P. Medlin, Steven L. Kronick, Karandeep Singh, Pauline K. Park, Lena M. Napolitano, Robert P. Dickson, Michael R. Mathis, Michael W. Sjoding, Andrew J. Admon, Kevin R. Ward, Christopher E. Gillies

**Affiliations:** Department of Emergency Medicine, University of Michigan, Ann Arbor; Department of Internal Medicine, University of Michigan, Ann Arbor; Department of Learning Health Sciences, University of Michigan, Ann Arbor; Department of Surgery, University of Michigan, Ann Arbor; Department of Microbiology & Immunology, University of Michigan, Ann Arbor; Department of Anesthesiology, University of Michigan, Ann Arbor; Department of Biomedical Engineering, University of Michigan, Ann Arbor; Michigan Institute for Data Science (MIDAS), University of Michigan, Ann Arbor; Michigan Center for Integrative Research in Critical Care (MCIRCC) University of Michigan, Ann Arbor

**Author notes:** **Corresponding Authors** Brandon C. Cummings, Christopher E. Gillies, 2800 Plymouth Road, NCRC Building 10, Ann Arbor, Michigan, 48109. These two authors have contributed equally to the work (Brandon C. Cummings, Sardar Ansari). **Conflicts of Interest and Source of Funding:** Christopher E. Gillies, Richard P. Medlin and Kevin R. Ward submitted a patent regarding our machine learning methodologies presented in this paper through the University of Michigan’s Office of Technology Transfer. Funded in part by the Michigan Institute for Data Science (MIDAS) “Propelling Original Data Science (PODS) Mini-Grants for COVID-19 Research” award.

## Abstract

**Introduction:** The 2019 coronavirus (COVID-19) has led to unprecedented strain on healthcare facilities across the United States. Accurately identifying patients at an increased risk of deterioration may help hospitals manage their resources while improving the quality of patient care. Here we present the results of an analytical model, PICTURE (**P**redicting **I**ntensive **C**are **T**ransfers and Other **U**nfo**R**eseen **E**vents), to identify patients at a high risk for imminent intensive care unit (ICU) transfer, respiratory failure, or death with the intention to improve prediction of deterioration due to COVID-19. We compare PICTURE to the Epic Deterioration Index (EDI), a widespread system which has recently been assessed for use to triage COVID-19 patients.

**Methods:** The PICTURE model was trained and validated on a cohort of hospitalized non-COVID-19 patients using electronic health record data from 2014-2018. It was then applied to two hold-out test sets: non-COVID-19 patients from 2019 and patients testing positive for COVID-19 in 2020. PICTURE results were aligned to the EDI for head-to-head comparison via Area Under the Receiver Operator Curve (AUROC) and Area Under the Precision Recall Curve (AUPRC). We compared the models’ ability to predict an adverse event (defined as ICU transfer, mechanical ventilation use, or death) at two levels of granularity: (1) maximum score across an encounter with a minimum lead time before the first adverse event and (2) predictions at every observation with instances in the last 24 hours before the adverse event labeled as positive. PICTURE and the EDI were also compared on the encounter level using different lead times extending out to 24 hours. Shapley values were used to provide explanations for PICTURE predictions.

**Results:** PICTURE successfully delineated between high- and low-risk patients and consistently outperformed the EDI in both of our cohorts. In non-COVID-19 patients, PICTURE achieved an AUROC (95% CI) of 0.819 (0.805 - 0.834) and AUPRC of 0.109 (0.089 - 0.125) on the observation level, compared to the EDI AUROC of 0.762 (0.746 - 0.780) and AUPRC of 0.077 (0.062 - 0.090). On COVID-19 positive patients, PICTURE achieved an AUROC of 0.828 (0.794 - 0.869) and AUPRC of 0.160 (0.089 - 0.199), while the EDI scored an AUROC of 0.792 (0.754 - 0.835) and AUPRC of 0.131 (0.092 - 0.159). The most important variables influencing PICTURE predictions in the COVID-19 cohort were a rapid respiratory rate, a high level of oxygen support, low oxygen saturation, and impaired mental status (Glasgow coma score).

**Conclusion:** The PICTURE model is more accurate in predicting adverse patient outcomes for both general ward patients and COVID-19 positive patients in our cohorts compared to the EDI. The ability to consistently anticipate these events may be especially valuable when considering a potential incipient second wave of COVID-19 infections. PICTURE also has the ability to explain individual predictions to clinicians by ranking the most important features for a prediction. The generalizability of the model will require testing in other health care systems for validation.

## 1. Introduction

The effect of the 2019 coronavirus (COVID-19) on the US healthcare system cannot be overstated. It has led to unprecedented clinical strain in hospitals across the nation, prompting the proliferation of ICU capability and of lower-acuity field hospitals to accommodate the increased patient load. A predictive early warning system capable of identifying patients at increased risk of deterioration could assist hospitals in maintaining a high level of patient care while more efficiently distributing their thinly stretched resources. However, a recent review has illustrated that high-quality, validated models of deterioration in COVID-19 patients are lacking [1]. All 16 of the models appraised in this review were rated at high or unclear risk of bias, mostly because of non-representative selection of control patients. A primary concern is that these models may overfit to the small COVID-19 datasets that are currently available.

Early warning systems have been and continue to be applied in hospital settings prior to the COVID-19 pandemic to predict patient deterioration events before they occur, giving healthcare providers time to intervene [2]. The prediction of adverse events such as ICU admission and death provides crucial information to avert impending critical deterioration: it is estimated that 85% of such events are preceded by detectable changes in physiological signs [3] that may occur up to 48 hours before the event [4]. In addition, approximately 44% of events are avoidable through early intervention, [5] and 90% of unplanned transfers to the intensive care unit (ICU) are preceded by a new or worsening condition [6], [7]. Such abnormal signals indicate that predictive data analytics may be used to alert providers of incipient deterioration events, ultimately leading to improved care and reduced costs [8], [9]. Given the number of unknowns surrounding the pathophysiology of COVID-19, early warning systems may play a pivotal role in treating patients and improving outcomes.

One model which has been assessed in COVID-19 patients is the Epic Deterioration Index (EDI) [10], [11]. The EDI has the advantage over models built on COVID-19 specific data in that it is not overfit to small datasets as it was trained on over 130,000 encounters [11], [12]. Recent work has suggested it may be capable of stratifying COVID-19 patients according to their risk of deterioration [11]. The outcomes used in this study were those considered most relevant to the care of COVID-19 patients including ICU level of care, mechanical ventilation, and death. While the EDI was able to successfully isolate groups of patients at very high and very low risk of deterioration, the overall performance as a continuous predictor was moderately low (AUROC 0.76 (95% CI 0.68-0.84), n = 174) [11]. Additionally, much of the detail surrounding the EDI’s structure and internal validation has not been shared publicly. This makes the interpretation of individual predictions difficult.

In this current report we have applied our previously described model, PICTURE (**P**redicting **I**ntensive **C**are **T**ransfers and other **U**nfo**R**eseen **E**vents) to a cohort of patients testing positive for COVID-19 [13]. Initially developed to predict patient deterioration in the general wards, we have re-trained the model to target those outcomes considered most relevant to the COVID-19 pandemic: ICU level of care, mechanical ventilation, and death. PICTURE, like the EDI, was trained and tuned on a large non-COVID-19 cohort (128,732 encounters). Furthermore, we took extensive steps in the PICTURE framework to limit overfitting and learning missingness patterns in the data. This is critical in providing clinicians with novel, useful, and generalizable alerts as missing patterns can vary in different settings and different patient phenotypes [13]. In addition to the risk score, PICTURE also provides actionable explanations for its predictions in the form of Shapley values which may help clinicians easily interpret scores and better determine if actionability on the alert is required [14]. We validate this system in both a non-COVID-19 cohort as well as in those testing positive for COVID-19, and compare it to the EDI on the same matched cohorts.

## 2. Methods

### 2.1 Setting and study population

The study protocol was approved by the University of Michigan’s Institutional Review Board (HUM00092309). EHR data was collected from a large tertiary, academic medical system (Michigan Medicine) from January 1, 2014 to June 1, 2020. The first five years of data (2014 – 2018, n = 128,732 encounters) were used to train and validate the model while 2019 data was reserved as a hold-out test set (n = 32,754 encounters). Training, validation, and test populations were segmented to prevent overlap of multiple hospital encounters between sets. Criteria for inclusion in these three cohorts were defined as age ≥ 18 and ≤ 89 years who were hospitalized (having inpatient or other observation status) in a general ward. We excluded patients with a left ventricular assist device (LVAD), patients who were discharged to hospice, and patients whose ICU transfer was from a floor other than a general ward (e.g. operating or interventional radiology unit) in order to exclude planned ICU transfers.

To be included in the COVID-19 cohort (n=430), patients must have received a positive COVID-19 test result from Michigan Medicine within a span of 14 days before or 21 days after their hospitalization. These patients were then filtered using the same criteria used in the 2019 test set, with the exception of the hospice and LVAD distinctions. Only discharged patients or those who already experienced an adverse event were included. **Table 1** describes the study cohort and the frequency of individual adverse events. When compared to the non-COVID-19 test cohort from 2019, the median age of COVID-19 patients was slightly higher (64.8 vs. 61.3) and the proportion of Black and Asian patients considerably higher. The rate of adverse events was also higher, rising from slightly under 4% to 30%.

**Table 1:** Study population. Patients were subset into one of four study cohorts: a training set for learning model parameters; a validation set for model structure and hyperparameter tuning; a hold-out test set for evaluation; and a final test set composed of patients testing positive for COVID-19. Values are based on individual hospital encounters. Age is presented as median with interquartile range (IQR). In our population, the three most frequently encountered races were White, Black, and Asian. Other races comprising less than 1% of the population each were incorporated under the “Other” heading. Sex is reported as the number and percent of females. The event rate represents a composite outcome indicating that one of the following events occurred: death, ICU transfer, mechanical ventilation, and cardiac arrest. The individual frequencies of these adverse events are also reported, and represent the number of cases where each particular outcome was the first to occur. Cardiac arrest was not used as a target in the COVID-19 positive population as the manually adjudicated data is not yet available. Please see Section 2.3 “Outcomes” for the procedure of calculating these targets.

| Dataset | Non-COVID-19 |  |  | COVID-19 |
| --- | --- | --- | --- | --- |
|  | Training<br>2014 – 2018 | Validation<br>2014 – 2018 | Test<br>2019 | Test<br>2020 |
| **n (encounters)** | 103,322 | 25,410 | 32,754 | 430 |
| **n (patients)** | 61,210 | 15,302 | 22,826 | 401 |
| **Age** median (IQR) | 59.7 (46.2, 70.0) | 59.9 (46.0, 70.5) | 61.30 (46.5, 70.80) | 63.1 (51.6, 72.0) |
| **Race** n (%) |  |  |  |  |
| White | 84,931 (82.20%) | 20,751 (81.66%) | 26,395 (80.59%) | 177 (41.16%) |
| Black | 11,965 (11.58%) | 3,030 (11.92%) | 4,171 (12.73%) | 182 (42.33%) |
| Asian | 2,051 (1.99%) | 524 (2.06%) | 667 (2.04%) | 26 (6.05%) |
| Other | 4,375 (4.23%) | 1,105 (4.35%) | 1,521 (4.64%) | 45 (10.47%) |
| **Female sex** n (%) | 51,886 (50.22%) | 12,715 (50.04%) | 16,336 (49.87%) | 196 (45.58%) |
| **Event rate** n (%) | 4,104 (3.97%) | 960 (3.78%) | 1,297 (3.96%) | 129 (30.00%) |
| Death | 867 (0.84%) | 188 (0.74%) | 255 (0.78%) | 11 (2.56%) |
| ICU transfer | 2,913 (2.82%) | 703 (2.77%) | 983 (3.00%) | 117 (27.21%) |
| Mechanical Ventilation | 1,311 (1.27%) | 300 (1.18%) | 348 (1.06%) | 46 (10.70%) |
| Cardiac arrest | 147 (0.14%) | 29 (0.11%) | 56 (0.17%) | N/A |

### 2.2. Predictors

The variables used as predictors were collected from the electronic health record (EHR) and broadly included vital signs and physiologic observations, laboratory and metabolic values, and demographics. Specific features were selected based on previous analysis [13]. Vital signs used in the model included heart rate, respiratory rate, pulse oximetry, Glasgow Coma Score (GCS), urine output, and blood pressure. Laboratory and metabolic features included electrolyte concentrations, glucose and lactate, and blood cell counts. Demographics include age, height, weight, race, and gender. Fluid bolus and oxygen supplementation were also included as features. A full list of features are presented in **Table S.1** in the supplemental material alongside their respective mean, standard deviation, and missingness rates.

### 2.3 Outcomes

The primary outcomes in the training, validation, and test cohorts (data collected from 2014 through 2019) were death, cardiac arrest (as defined by the American Heart Association’s *Get With The Guidelines*^®^), transfer to an ICU from a general ward or similar unit, or need for mechanical ventilation. Determination of ICU transfer was based on actual location or accommodation level. Outcomes in the COVID-19 positive cohort differed slightly in two respects. First, cardiac arrest information was not available at the time of writing and so was not included. Second, the emergency procedures undertaken by the hospital to accommodate the high volume of COVID-19 patients led to the delivery of critical care in non-ICU settings. Thus, ICU level of care was used to determine ICU transfer rather than actual location. In other words, implementation of ICU care that otherwise would not have been provided to patients in a non-ICU setting but was provided due to capacity constraints was determined to be ICU level care or ICU transfer. Observations occurring thirty minutes before the first event or later were discarded to be consistent with other approaches [15]. For observation level predictions, individual observations were labeled positive if they occurred within 24 hours of any of the above events, and negative otherwise. We refer to these composite adverse events as the “outcome” or “target” throughout the text. These outcomes were designed to closely follow those of a recent analysis of the EDI at Michigan Medicine [11].

### 2.4 PICTURE model development

To train and evaluate the PICTURE model, we partitioned our data into four folds: a training and validation set using data from 2014 - 2018, a test set using 2019 data, and a fourth set consisting of data from COVID-19 positive patients. The sets were partitioned such that multiple hospital encounters from the same individual were restricted to one cohort, preventing patient-level overlap between cohorts. Encounters with an admission date from January 1, 2014 to December 31, 2018 were used for training and validation/hyperparameter tuning (n = 128,732 encounters). These patients were further divided between training and validation sets using an 80/20% split. Those patients with an admission date between January 1, 2019 and December 31, 2019 were reserved as a hold-out test set (n = 32,754 encounters). Lastly, patients testing positive for COVID-19 from March 1, 2020 to June 1, 2020 were reserved as a separate set (n = 430 encounters).

The training and validation sets were grouped into 8-hour windows to ensure that each encounter would have the same amount of observations for the same amount of time in the hospital, avoiding emphasis on patients who get more frequent updates while training the model. The 2019 and COVID-19 test sets were left in a granular format, where each new observation represented the addition of new data (e.g. an updated vital sign). Vital signs and laboratory values were forward filled such that each observation represented the most up-to-date information available as of that time, and the remaining missing values were iteratively imputed using the mean of the posterior distribution from a multivariate Bayesian regression model. This method has previously been demonstrated to reduce the degree to which tree-based models learn missingness patterns in order to bolster performance [13]. Classification was achieved using an XGBoost model (v. 0.90) using a logistic objective function with a maximum tree depth of three nodes and stopped when the validation AUPRC had not improved for 30 rounds [16]. All analysis was performed using Python 3.8.2.

### 2.5 Epic deterioration index

The EDI is a proprietary model developed by Epic Systems Corporation (Verona, WI). Michigan Medicine utilizes EPIC as its electronic medical record system and has access to the EDI tool. Similar to PICTURE, it uses clinical data that are commonly available in the EHR to make predictions regarding patient deterioration. It was trained using a similar composite outcome as a target including death, rapid response team (RRT) call, ICU transfer, and resuscitation as adverse events [11]. It is calculated every 15 minutes. Specific details surrounding its structure, parameters, or training procedures have not been shared publicly.

### 2.6 PICTURE model evaluation

#### 2.6.1 Evaluation of PICTURE performance in non-COVID-19 cohort

The performance of the PICTURE model was first assessed on all 32,754 encounters in the hold-out test set comprising patients from 2019. Another early warning aggregate score, National Early Warning Score (NEWS), was used for comparison in this preliminary analysis [17], [18]. The original NEWS was selected over the updated NEWS2 score due to evidence that its performance was found to be higher when predicting adverse events in patients at risk of respiratory failure [19]. For each observation time point, the NEWS score was calculated according to their published scoring system and compared to PICTURE scores. Performance was assessed on two scales: observation-level and encounter-level. The term “observation-level” is used to denote the performance of the model at each time the data for a patient is updated, with observations occurring 24 hours prior to a target event marked as 1 and otherwise marked as 0. “Encounter-level” describes the model performance across the entire hospital encounter for one patient, and refers to the maximum model score during the patient’s stay and occuring at least 30 minutes (or longer for different minimal lead times) before the first event. The target in this case is a one if the patient ever met an outcome condition during their stay, and zero otherwise.

#### 2.6.2 Comparison of PICTURE to EDI

Since the EDI makes a prediction every 15 minutes, we were not able to calculate scores at each timepoint as with NEWS. We simulated how the PICTURE score, calculated at irregular intervals each time a new data point arrives, would align with the EDI scores calculated every 15 minutes. This limited the available number of encounters to 21,215 encounters in the 2019 test set, and 401 encounters in the COVID-19 cohort. The PICTURE scores were merged onto EDI values by taking the most recent PICTURE prediction before the EDI prediction. This was to give the EDI any advantages in the alignment procedure. **Figure 1** displays a visual schematic of this alignment. The two models were then evaluated using the same observation- and encounter-level methods described in the previous section.

**Figure 1:**
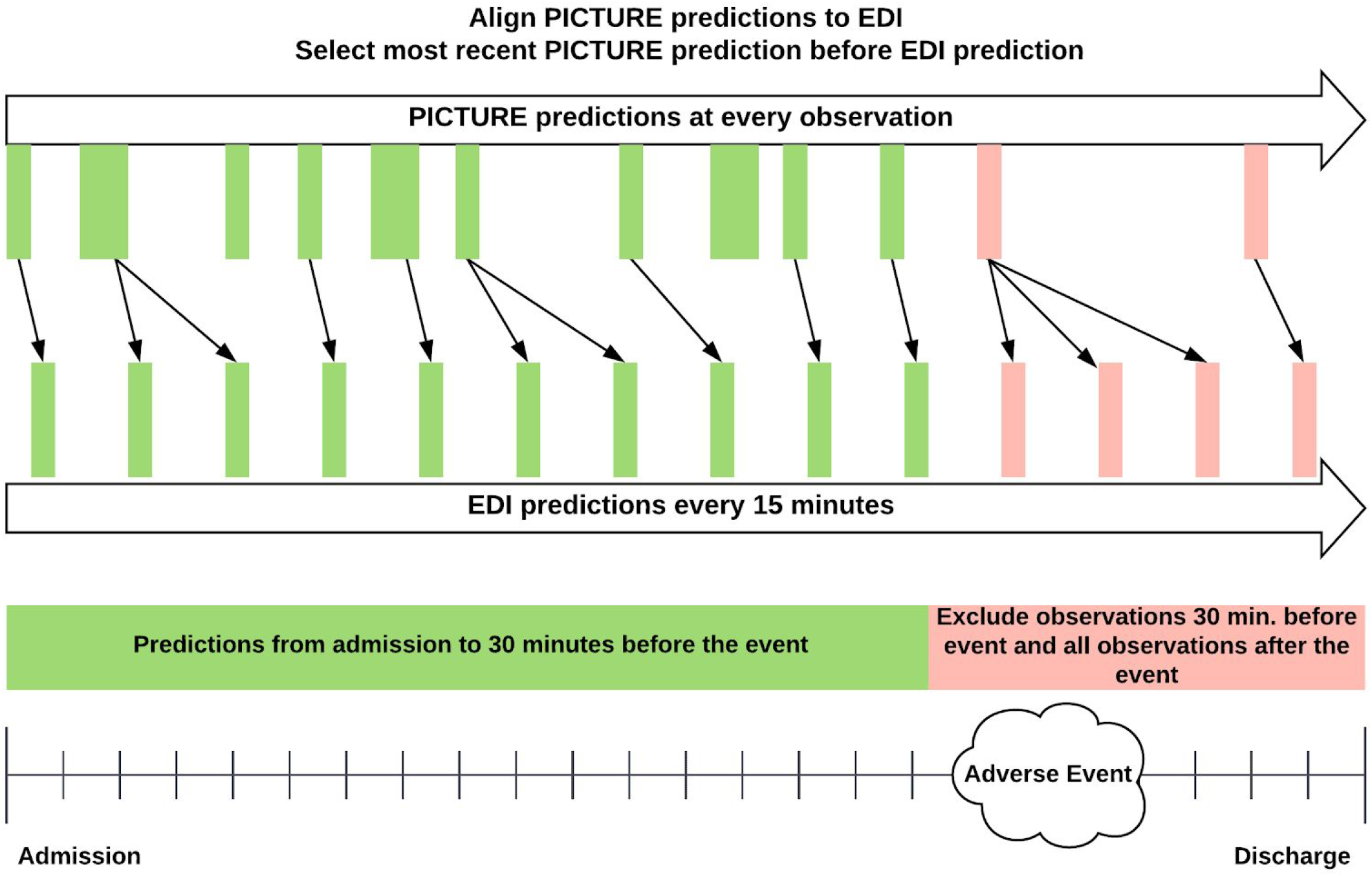
Alignment of PICTURE predictions to EDI scores. While the PICTURE system outputs predictions each time a new observation (e.g. a new vital sign) is input into the system, the EDI score is generated every 15 minutes. To give the EDI any potential advantage, PICTURE scores are aligned to EDI scores by selecting the most recent PICTURE score before each EDI prediction. In both cases, observations occurring 30 minutes before the target and after are excluded (red). For the patients who did not experience an adverse event, the maximum score was calculated across the entire encounter.

#### 2.6.3 Performance measures

Area under the Receiver Operating characteristic curve (AUROC) and area under the precision-recall curve (AUPRC) were used as the primary criteria for comparison between the models. AUROC can be interpreted as the probability two randomly chosen observations (one with a positive target, the other negative) are ranked in the correct order by the model prediction score. AUPRC describes the average positive predictive value (PPV) across the range of sensitivities. 95% confidence intervals were calculated for encounter-level statistics with a bootstrap method using 1,000 replications to compute pivotal confidence intervals. For observation-level statistics, block bootstrapping was used to ensure randomization between encounters and within the observations of an encounter. P-values were calculated using a generalized two-sample, one-tailed t-test.

#### 2.6.4 Feature ranking and prediction explanation

Despite the many benefits yielded by increasingly advanced machine learning models, use of these models in the medical field has lagged behind other fields. One contributing factor is the complexity of these models, which make the resulting predictions difficult to interpret and in turn make it difficult to build clinician trust [20]. To better provide insight into the PICTURE predictions, tree-based Shapley values are calculated for each observation. Borrowed from game theory, Shapley values describe the relative contribution of a feature to the model’s prediction [14], [21]. Positive values denote features that influenced the model toward a high prediction score (here indicating a higher likelihood of an adverse event), while negative values indicate the feature pushed the model toward a lower prediction score. The sum of the Shapley values across a single prediction plus the mean log-odds probability of the model is equal to the log-odds of the prediction probability. Shapley values can be used to provide insight into individual model predictions or aggregated to visualize global variable importance.

## 3. Results and Discussion

### 3.1 Evaluation of PICTURE performance in non-COVID-19 cohort

The ability of the PICTURE model to accurately predict the composite target was first assessed using the 32,754 encounters in the hold-out test set from 2019. To provide a baseline for comparison, National Early Warning Scores (NEWS) were calculated alongside each PICTURE prediction output. The observation- and encounter-level AUROC and AUPRC are presented with 95% confidence intervals in **Table 2** below. The observation-level event rate can be interpreted as the fraction of individual observations during which an adverse event occurred within 24 hours, while the encounter-level event rate refers to the proportion of hospital encounters experiencing such an event. The difference in AUROC between PICTURE and NEWS was 0.068 (95% CI: 0.057 - 0.081) on the observation level and 0.093 (95% CI: 0.083 - 0.102) on the encounter level, indicating statistical significance at ɑ = 0.05 as the confidence interval does not include zero. The difference in AUPRC was similarly significant, at 0.045 (95% CI: 0.034 - 0.054) and 0.181 (95% CI: 0.157 - 0.201) on the observation and encounter level respectively.

**Table 2:** Evaluation of PICTURE performance in a non-COVID-19 cohort. Area under the Receiver Operator characteristic curve (AUROC), area under the precision-recall curve (AUPRC), and the event rate are presented on the observation level as well as aggregated across each hospital encounter. 95% confidence intervals were calculated using a block bootstrap with 1000 replicates. In the case of the observation-level, this bootstrap was blocked on the encounter level. The National Early Warning System (NEWS) score is used as a baseline for comparison. n = 32,754 encounters. * = p-value < 0.05, ** = p-value < 0.001.

| Analytic | Granularity | AUROC (95% CI) | AUPRC (95% CI) | Event rate (%) |
| --- | --- | --- | --- | --- |
| PICTURE | Observation-level | 0.823 (0.812 - 0.836)** | 0.099 (0.085 - 0.112)** | 1.01% |
| NEWS | Observation-level | 0.754 (0.742 - 0.767)** | 0.054 (0.048 - 0.060)** |  |
| PICTURE | Encounter-level | 0.850 (0.837 - 0.862)** | 0.318 (0.291 - 0.342)** | 3.96% |
| NEWS | Encounter-level | 0.757 (0.744 - 0.770)** | 0.137 (0.121 - 0.151)** |  |

### 3.2 Comparison of PICTURE to EDI in a non-COVID-19 cohort

PICTURE was then compared to the EDI model on non-COVID patients in the same hold-out test set from 2019. Due to limitations in available EDI scores, the number of encounters was restricted to 21,215. These time-matched scores were again evaluated using AUROC and AUPRC on the observation- and encounter levels (**Table 3**). **Figure 2** displays the associated ROC and PR curves for the observation-level performance. The difference of AUROC and AUPR between PICTURE and the EDI reached statistical significance (α = 5%) on both the observation level (AUROC: 0.057 [95% CI: 0.044 - 0.069], AUPRC: 0.032 [95% CI: 0.019 - 0.043]) and the encounter level (AUROC: 0.059 [95% CI: 0.048 - 0.069], AUPRC: 0.095 [95% CI: 0.067 - 0.118]).

**Table 3:** Comparison of PICTURE and the EDI in a non-COVID-19 cohort. Area under the Receiver Operator Characteristic curve (AUROC), area under the precision-recall curve (AUPRC), and the event rate are presented on the observation level as well as aggregated across each hospital encounter. n = 21,215 encounters. * = p-value < 0.05, ** = p-value < 0.001.

| Analytic | Granularity | AUROC (95% CI) | AUPRC (95% CI) | Event rate (%) |
| --- | --- | --- | --- | --- |
| PICTURE | Observation-level | 0.819 (0.805 - 0.834)** | 0.109 (0.089 - 0.125)** | 0.75% |
| EDI | Observation-level | 0.762 (0.746 - 0.780)** | 0.077 (0.062 - 0.090)** |  |
| PICTURE | Encounter-level | 0.862 (0.850 - 0.876)** | 0.362 (0.327 - 0.394)** | 4.17% |
| EDI | Encounter-level | 0.803 (0.787 - 0.821)** | 0.267 (0.240 - 0.295)** |  |

**Figure 2:**
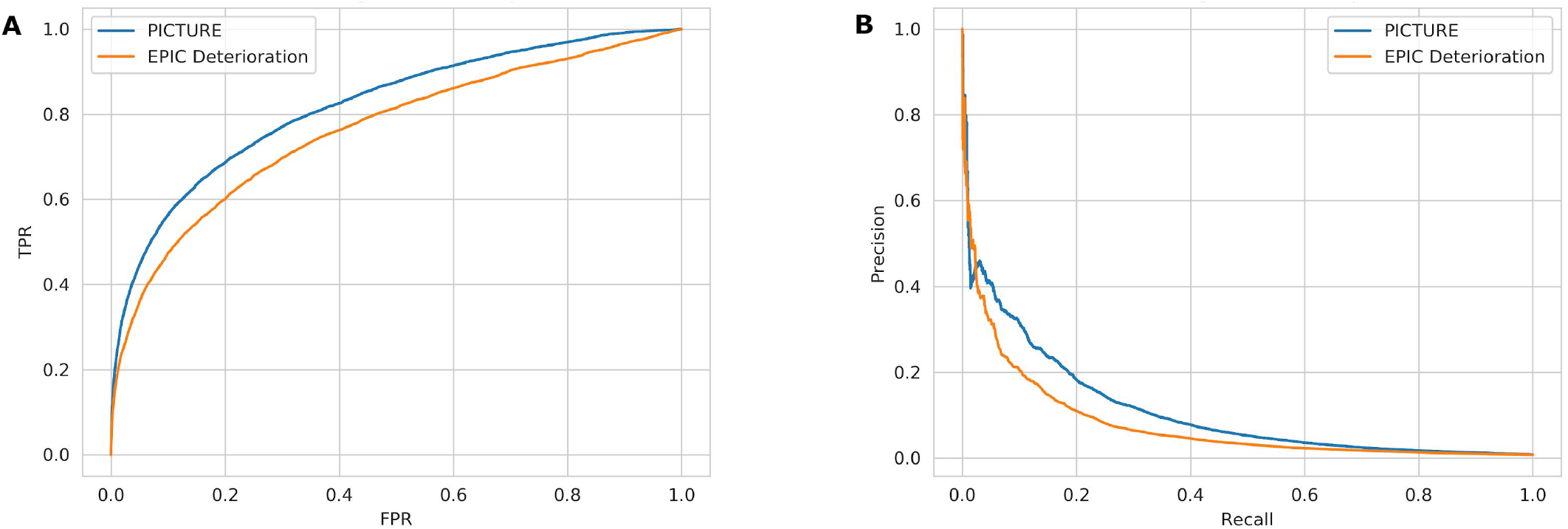
Comparison of PICTURE and the EDI in a non-COVID-19 cohort. Panel A: Receiver Operating Characteristic (ROC) curves for PICTURE and EDI models. PICTURE AUC: 0.819. EDI AUC: 0.762. Panel B: Precision-recall (PR) curves for the two models. PICTURE AUC: 0.109. EDI AUC: 0.077. Both curves represent observation-level analysis.

In addition to classification performance, lead time represents another critical component of a predictive analytics’ utility as it determines the amount of time clinicians have to act on the model’s recommendations. We assessed the model’s relative performance at different lead times in a threshold-independent manner by censoring data occurring 0.5, 1, 2, 6, 12, and 24 hours before an adverse event (**Table 4**). In our cohort, PICTURE performs markedly better than the EDI model even when considering predictions made 24 hours or more before the actual event.

**Table 4:** Lead time analysis in non-COVID-19 cohort. The performance of the two models (encounter-level) at various lead times were assessed by evaluating the maximum prediction score prior to *x* hours before the given event, with *x* ranging in progressively greater intervals from 0.5 to 24. On this cohort of non-COVID-19 subjects, PICTURE consistently outperformed the EDI model at each level of censoring.

| Lead time (hours) | AUROC |  | AUPRC |  | Event Rate (%) | Sample size (n encounters) |
| --- | --- | --- | --- | --- | --- | --- |
|  | PICTURE | EDI | PICTURE | EDI |  |  |
| 0.5 | 0.862 | 0.803 | 0.365 | 0.268 | 4.17% | 21215 |
| 1 | 0.853 | 0.795 | 0.343 | 0.248 | 4.15% | 21211 |
| 2 | 0.840 | 0.784 | 0.315 | 0.232 | 4.11% | 21201 |
| 6 | 0.827 | 0.766 | 0.276 | 0.204 | 3.89% | 21153 |
| 12 | 0.818 | 0.765 | 0.243 | 0.176 | 3.63% | 21096 |
| 24 | 0.809 | 0.757 | 0.200 | 0.139 | 3.20% | 21003 |

### 3.3 Comparison of PICTURE to EDI in COVID-19 patients

When applied to patients testing positive for COVID-19, PICTURE performs similarly well. PICTURE scores were again aligned to EDI scores using the process outlined in Section 2.6.2. This resulted in the inclusion of 402 encounters. **Table 5** presents AUROC and AUPRC values for PICTURE and EDI on both the observation and encounter level with 95% confidence intervals, and **Figure 3** displays the associated ROC and PR curves. On the observation level, the difference in AUROC between PICTURE and the EDI reached statistical significance (α = 5%) (0.037 [95% CI: 0.006 - 0.067]) but the difference in AUPRC (0.028 [95% CI: −0.018 - 0.058]) did not reach significance. Both the AUROC and AUPRC differences (95% CI) reached significance on the encounter level (AUROC: 0.102 [0.064 - 0.1381], AUPRC: 0.124 [0.048 - 0.179]). Of note, the EDI results at the observation-level (AUROC 0.792 [0.754 - 0.835]) were very similar to those described in a previous validation (AUROC 0.76 [95% CI 0.68-0.84]) though with a smaller confidence interval due to a larger sample size [11].

**Table 5:** Comparison of PICTURE and the EDI in patients testing positive for COVID-19. Area under the Receiver Operator Characteristic curve (AUROC), area under the precision-recall curve (AUPRC), and the event rate are presented on the observation level as well as aggregated across each hospital encounter. n = 401. * = p-value < 0.05, ** = p-value < 0.001.

| Analytic | Granularity | AUROC (95% CI) | AUPRC (95% CI) | Event rate (%) |
| --- | --- | --- | --- | --- |
| PICTURE | Observation-level | 0.828 (0.794 - 0.869)* | 0.160 (0.089 - 0.199) | 3.75% |
| EDI | Observation-level | 0.792 (0.754 - 0.835)* | 0.131 (0.092 - 0.159) |  |
| PICTURE | Encounter-level | 0.876 (0.839 - 0.913)** | 0.664 (0.571 - 0.739)** | 24.9% |
| EDI | Encounter-level | 0.774 (0.721 - 0.826)** | 0.541 (0.458 - 0.623)** |  |

**Figure 3:**
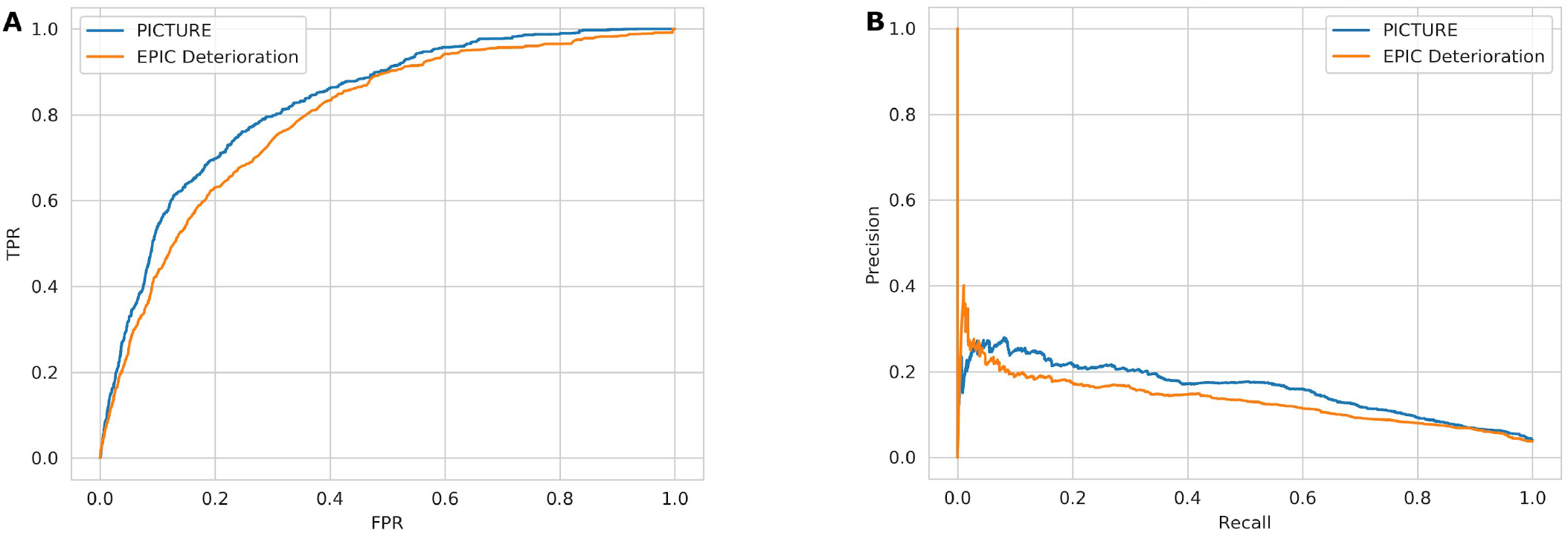
Comparison of PICTURE and the EDI in patients testing positive for COVID-19. Panel A: Receiver Operator Characteristic (ROC) curves for PICTURE and EDI models. PICTURE AUC: 0.828. EDI AUC: 0.792. Panel B: Precision-recall (PR) curves for the two models. PICTURE AUC: 0.160. EDI AUC: 0.131. Both curves represent observation-level analysis.

As with the non-COVID-19 cohort, a similar lead time analysis was then performed to assess the performance of PICTURE and EDI when making predictions further in advance. Thresholds were again set at 0.5, 1, 2, 6, 12, and 24 hours before the event and observations occurring after this cutoff were censored. In our cohort, PICTURE again out-performed the EDI even when making predictions 24 hours in advance (**Table 6**).

**Table 6:** Lead time analysis in COVID-19 cohort. The performance of the two models (encounter-level) at various lead times were again assessed by evaluating the maximum prediction score prior to *x* hours before the given event, with *x* ranging in progressively greater intervals from 0.5 to 24. On this cohort of COVID-19 subjects, PICTURE consistently outperformed the EDI model at each level of censoring.

| Lead time<br>(hours) | AUROC |  | AUPRC |  | Event Rate (%) | Sample size<br>(n encounters) |
| --- | --- | --- | --- | --- | --- | --- |
|  | PICTURE | EDI | PICTURE | EDI |  |  |
| **0.5** | 0.876 | 0.774 | 0.683 | 0.551 | 24.94% | 401 |
| **1** | 0.865 | 0.767 | 0.648 | 0.532 | 24.75% | 400 |
| **2** | 0.849 | 0.760 | 0.618 | 0.517 | 24.37% | 398 |
| **6** | 0.826 | 0.746 | 0.569 | 0.482 | 23.02% | 391 |
| **12** | 0.802 | 0.731 | 0.527 | 0.447 | 21.00% | 382 |
| **24** | 0.787 | 0.721 | 0.472 | 0.413 | 18.87% | 371 |

### 3.4 Explanations of predictions

To provide clinicians with a description of factors influencing a given PICTURE score, we used Shapley values computed at each observation. **Figure 4** depicts an aggregated summary of the 20 most influential features in the 2019 test set (Panel A) and in the COVID-19 set (Panel B). Positive Shapley values indicate that the variable pushed the PICTURE score toward a positive decision (i.e. predicting an adverse event). While many of the feature rankings appear similar between the 2019 and COVID-19 cohorts, we noted that respiratory variables such as respiratory rate, oxygen support, and SpO_2_ played a more pronounced role in predicting adverse events in COVID-19 positive patients than in non-COVID-19 patients. One point of note is that the amount of oxygen support played a significant role in both cohorts. While the EDI does not use the amount of oxygen support as a continuous variable, it does have a feature termed “oxygen requirement” [11]. To demonstrate that the observed improvement of PICTURE over the EDI is not driven solely by this additional information, oxygen support was binarized and the PICTURE model retrained. While performance did decrease, indicating that the inclusion of oxygen support as a continuous variable is useful in predicting deterioration, PICTURE still outperformed the EDI on both the non-COVID-19 (difference in AUROC: 0.031, AUPRC: 0.044) and COVID-19 (difference in AUROC: 0.036, AUPRC: 0.019) cohorts.

**Figure 4:**
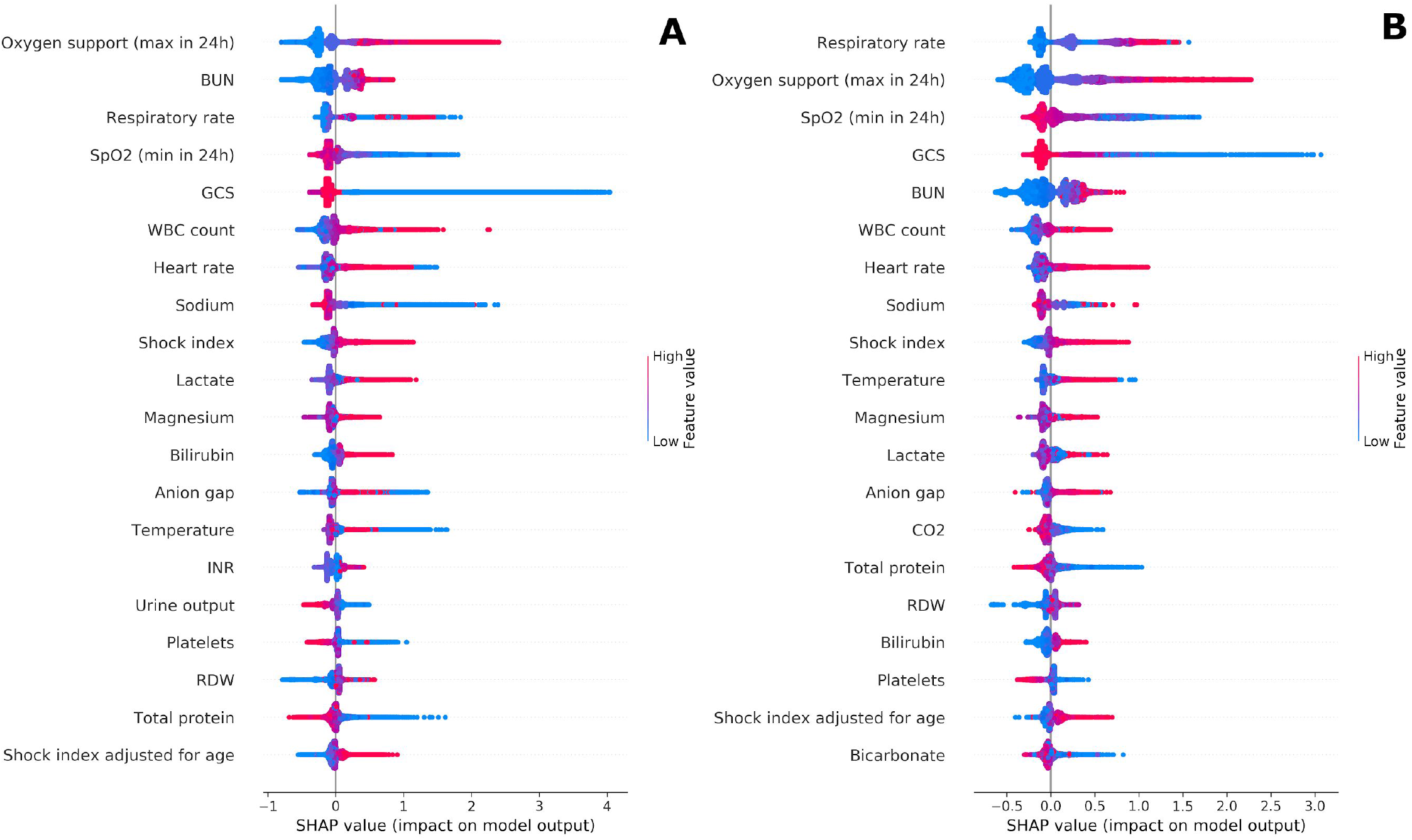
Shapley summary plots. Panel A) depicts an aggregated summary plot of the Shapley values from the 2019 test set, while Panel B) corresponds to COVID-19 positive patients. The 20 most influential features are ranked from top to bottom, and the distribution of Shapley values across all predictions are plotted. The magnitude of the Shapley value is displayed on the horizontal axis, while the value of the feature itself is represented by color. For example, a large amount of oxygen support over 24 hours (red) in Panel A was associated with a highly positive influence on the model, while low to no oxygen support (blue) pushed the model back towards 0. BUN = blood urea nitrogen; GCS = Glasgow Coma Score; INR = International Normalized Ratio; RDW = red cell distribution width; WBC = white blood cells.

### 3.5 Case study

As a demonstration of the potential utility of PICTURE, an individual hospital encounter was selected and the trajectories of PICTURE and the EDI are visualized in **Figure 5**. A previous study assessing the use of EDI in COVID-19 patients found that an EDI score of 64.8 or greater to be an actionable threshold to identify patients at increased risk [11]. As PICTURE scores lie on a different scale than the EDI, we determined two comparable thresholds using sensitivity and PPV on the COVID-19 cohort. Due to the high event rate in this cohort, alert thresholds in non-COVID-19 patients may be lower. At a threshold of 64.8, we found that the EDI had a sensitivity of 0.164, specificity of 0.971, positive predictive value (PPV) of 0.184, and negative predictive value (NPV) of 0.968. When aligned by sensitivity, a PICTURE threshold of 0.183 corresponded to a specificity of 0.975, PPV of 0.217, and NPV of 0.968. Likewise when aligned by PPV, a PICTURE threshold of 0.075 corresponded to a sensitivity of 0.353, a specificity of 0.940, and NPV of 0.974. These two thresholds are present in Panel A of **Figure 5** below, while the original threshold of 64.8 is depicted in Panel B.

**Figure 5:**
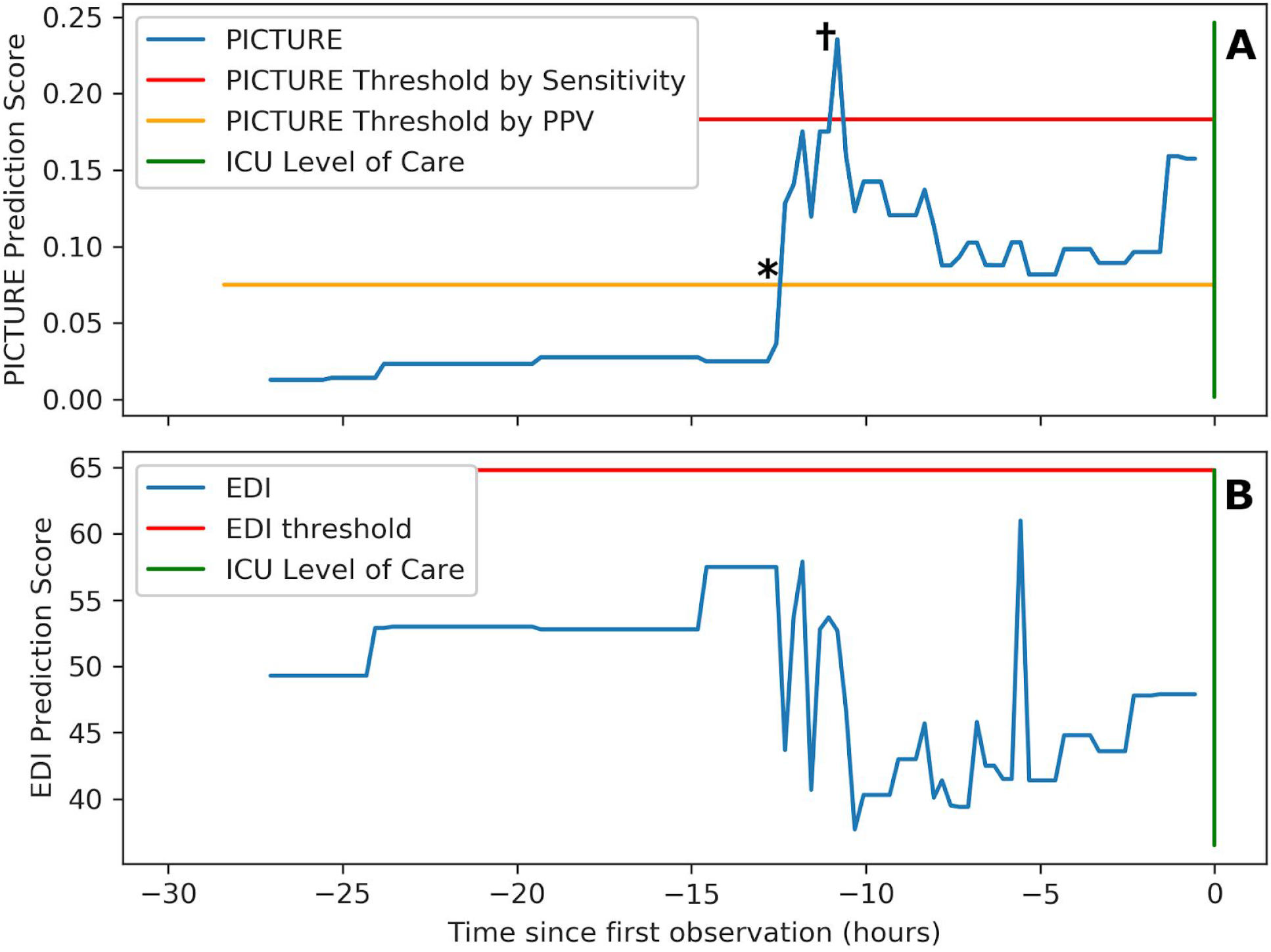
Sample trajectory of one patient. Panel A) depicts the PICTURE predictions over 27-hours before the patient is eventually transferred to an ICU level of care (green bar). Two possible alert thresholds are noted: one (red) based on the EDI’s sensitivity at a threshold of 64.8 (as suggested by [11]) while the other (yellow) is based on the EDI’s PPV at this threshold. Note that PICTURE peaks above the sensitivity-based threshold approximately 11 hours in advance of the ICU transfer, and then remains elevated over the PPV threshold until the transfer occurs. * and † represent the first time points at which PICTURE crossed each threshold, referenced in Table 7 below. Panel B) demonstrates the EDI over the same time range, with the threshold of 64.8 suggested by [11] in red.

Note that the PICTURE score remains low until approximately 12.5 hours before the adverse event (in this case, transfer to an ICU level of care), where it crosses the PPV-aligned threshold. Approximately 11 hours before the event, the PICTURE score peaks at a value of 0.235 and exceeds the sensitivity-aligned threshold of 0.18. After the initial peak, the PICTURE score then remains elevated, staying above the PPV-aligned threshold of 0.075 until the patient is transferred. In contrast, the EDI score never exceeded its alert threshold and it dropped when the PICTURE score increased

To simulate what a clinician receiving an alert from PICTURE might encounter, the Shapley values explaining the PICTURE predictions at both alert thresholds are recorded in **Table 7** below. Note that these features are dominated by respiratory features, though heart rate and temperature are also present. While these features may seem obvious in predicting the need for ICU care, it is worth highlighting that the EDI did not identify this patient as being at risk.

**Table 7:** Sample PICTURE explanations. The top 5 features corresponding to PICTURE predictions as it crosses the PPV-aligned threshold (*) and the sensitivity-aligned threshold (^†^) as noted in Figure 5. These predictions represent two possible locations where a clinician could receive an alert that their patient is deteriorating. For each prediction, the top 5 features as measured by Shapley values are reported. Such information could be shared alongside the prediction score to provide better clinical utility to healthcare providers. The number of standard deviations from the mean are included for comparison, and are calculated using the COVID-19 dataset. Note that oxygenation (supplemental oxygen, SpO_2_, and respiratory rate) and temperature play a dominant role in both cases. Heart rate represented the primary difference between these two timepoints. When the PICTURE score first exceeded the PPV threshold 12.5 hours before the ICU transfer, heart rate remained at 78 bpm and was not among the top features as measured by Shapley. At 11 hours before the event, when the PICTURE score was at its highest, heart rate had jumped to 124 bpm and was the fourth-most influential feature as measured by Shapley values.

| Shapley values after PPV threshold ( t -12.5h * ) |  |  |  |  |  |
| --- | --- | --- | --- | --- | --- |
| Rank | Feature name | Value | Mean (SD) | n Std. from Mean | Shapley Score |
| 1 | Oxygen supplementation (rolling 24h max) | 35.0 L/min | 3.98 (10.3) | 3.0 | + 1.97 |
| 2 | SpO<sub>2</sub> (rolling 24h min) | 85 % | 91.4 (3.65) | -1.7 | + 1.14 |
| 3 | Respiratory rate | 24 bpm | 20.8 (3.44) | 0.9 | + 0.77 |
| 4 | Temperature | 39.1 °C | 37.2 (0.43) | 4.5 | + 0.50 |
| 5 | Hematocrit | 32.3 | 37.3 (5.80) | -0.8 | + 0.09 |
| Shapley values after sensitivity threshold ( t -11h + ) |  |  |  |  |  |
| Rank | Feature name | Value | Mean (SD) | n Std. from Mean | Shapley Score |
| 1 | Oxygen supplementation (rolling 24h max) | 35 L/min | 3.98 (10.3) | 3.0 | + 2.00 |
| 2 | SpO<sub>2</sub> (rolling 24h min) | 85 % | 91.4 (3.65) | -1.7 | + 1.23 |
| 3 | Respiratory rate | 24 bpm | 20.8 (3.44) | 0.9 | + 0.74 |
| 4 | Heart rate | 124 bpm | 85.8 (13.6) | 2.8 | + 0.56 |
| 5 | Temperature | 39.1 °C | 37.2 (0.43) | 4.5 | + 0.46 |

## 4. Conclusion

The PICTURE early warning system accurately predicts adverse patient outcomes including ICU transfer, mechanical ventilation, and death at the Michigan Medicine. The ability to consistently anticipate these events may be especially valuable when considering a potential impending second wave of COVID-19 infections. The EDI is a widespread deterioration model which has recently been assessed in a COVID-19 population. Both PICTURE and the EDI were trained using approximately 130,000 encounters for general deterioration and thus are not overfit to the COVID-19 population [11], [12]. Using a head-to-head comparison, we demonstrated that PICTURE has higher performance than the EDI at a statistically significant level (α = 5%) for both COVID-19 positive and non-COVID-19 patients. In addition, PICTURE was capable of accurately predicting adverse events as far as 24 hours before the event occured. Lastly, PICTURE has the ability to explain individual predictions to clinicians by displaying those variables which most influenced its prediction using Shapley values. This analysis is limited to a single academic medical center, and its generalizability to other healthcare systems will require future study.

## Data Availability

Data and source code are not publicly available at this time.

## Supplemental material

**Table S.1:** Feature list with descriptive statistics. This table displays all features used in the PICTURE model along with their means, standard deviation, and missing rate in the training, validation, non-COVID-19 test, and COVID-19 cohorts. Values are computed on the encounter-level to avoid weighting the statistics by the number of observations per patients. For example, the mean value is first calculated for each patient, and these individual means are then averaged across all patients. Standard deviation was computed by again taking the mean value for each patient, and then calculating the standard deviation across all patients. Missingness rate represents the fraction of patients for whom there was no data available for the duration of their encounter. Gender and race are recorded as 1 if the patient meets the criteria (e.g. is female) and 0 otherwise. IV fluid bolus indicates a 1/0 flag if the patient received a fluid bolus during their stay. Oxygen supplementation represents the maximum amount of oxygen received by the patient in the last 24 hours, and zero if no oxygen order was placed. Pulse pressure is defined as diastolic blood pressure subtracted from systolic. Shock index is calculated as heart rate over systolic blood pressure, then multiplied by age in the case of the age-adjusted variable. SpO_2_ represents the minimum SpO_2_ in the last 24 hours. Abbreviations: BUN (blood urea nitrogen), GCS (Glasgow coma score), INR (international normalized ratio), MAP (mean arterial pressure), MCH (mean corpuscular hemoglobin), MCHC (mean corpuscular hemoglobin concentration), MCV (mean corpuscular volume), MPV (mean platelet volume), PT (prothrombin time), PTT (partial thromboplastin time), RBC (red blood cell count), RDW (red cell distribution width), SpO_2_ (peripheral oxygen saturation), WBC (white blood cell count).

| Feature Name | Training<br>(n=103,322 encounters) |  |  | Validation<br>(n = 25,410 encounters) |  |  | Test<br>(n = 32,754 encounters) |  |  | COVID-19<br>(n = 430 encounters) |  |  |
| --- | --- | --- | --- | --- | --- | --- | --- | --- | --- | --- | --- | --- |
|  | Mean | Std. dev. | Missing (%) | Mean | Std. dev. | Missing (%) | Mean | Std. dev. | Missing (%) | Mean | Std. dev. | Missing (%) |
| Age (years) | 57.35 | 17.14 | 0.0% | 57.54 | 17.38 | 0.0% | 57.78 | 17.17 | 0.0% | 61.84 | 15.05 | 0.0% |
| Albumin | 3.67 | 0.60 | 32.5% | 3.67 | 0.60 | 32.3% | 3.76 | 0.59 | 29.1% | 3.68 | 0.45 | 5.1% |
| Anion gap | 11.91 | 2.32 | 0.7% | 11.93 | 2.34 | 0.7% | 11.96 | 2.36 | 0.6% | 13.27 | 2.36 | 2.1% |
| Bicarbonate | 26.21 | 4.76 | 62.9% | 26.15 | 4.71 | 62.3% | 26.32 | 4.65 | 52.6% | 25.76 | 4.27 | 23.0% |
| Bilirubin | 1.03 | 2.36 | 34.3% | 0.98 | 2.17 | 34.3% | 0.99 | 2.16 | 30.3% | 0.65 | 0.52 | 5.3% |
| BUN | 19.77 | 14.39 | 0.8% | 19.85 | 14.05 | 0.9% | 21.07 | 15.28 | 0.7% | 23.63 | 20.00 | 2.6% |
| Calcium | 8.89 | 0.59 | 0.6% | 8.89 | 0.59 | 0.7% | 8.81 | 0.57 | 0.5% | 8.58 | 0.53 | 2.1% |
| Chloride | 104.66 | 4.22 | 0.6% | 104.69 | 4.17 | 0.6% | 104.58 | 4.22 | 0.5% | 102.84 | 4.56 | 1.2% |
| Chloride | 26.31 | 3.27 | 0.7% | 26.29 | 3.24 | 0.7% | 26.10 | 3.24 | 0.6% | 25.82 | 3.16 | 2.1% |
| Creatinine | 1.12 | 1.19 | 0.7% | 1.14 | 1.25 | 0.7% | 1.22 | 1.32 | 0.5% | 1.53 | 1.88 | 2.1% |
| Diastolic blood pressure | 68.81 | 9.38 | 0.1% | 69.05 | 9.30 | 0.0% | 68.73 | 10.02 | 0.0% | 70.23 | 8.92 | 0.5% |
| Gender (is Female) | 0.50 | 0.50 | 0.0% | 0.50 | 0.50 | 0.0% | 0.50 | 0.50 | 0.0% | 0.46 | 0.50 | 0.0% |
| GCS | 14.84 | 0.59 | 20.0% | 14.82 | 0.66 | 19.7% | 14.86 | 0.54 | 0.2% | 14.69 | 0.99 | 5.1% |
| Glucose | 125.18 | 42.21 | 0.5% | 125.71 | 42.44 | 0.5% | 125.52 | 43.50 | 0.3% | 125.27 | 51.77 | 1.2% |
| Height | 66.85 | 4.40 | 12.9% | 66.90 | 4.33 | 13.0% | 66.88 | 4.32 | 10.7% | 67.01 | 4.02 | 17.0% |
| Hematocrit | 34.77 | 6.12 | 0.6% | 34.85 | 6.09 | 0.6% | 35.13 | 6.33 | 0.5% | 37.27 | 5.80 | 1.9% |
| Hemoglobin | 11.54 | 2.22 | 0.6% | 11.57 | 2.21 | 0.7% | 11.45 | 2.23 | 0.5% | 12.11 | 2.11 | 1.9% |
| INR | 1.23 | 0.95 | 42.2% | 1.23 | 2.15 | 42.6% | 1.22 | 0.49 | 43.6% | 1.18 | 0.53 | 47.9% |
| IV fluid bolus | 0.68 | 0.39 | 0.0% | 0.68 | 0.39 | 0.0% | 0.59 | 0.41 | 0.0% | 0.41 | 0.42 | 0.0% |
| Lactate | 1.52 | 0.88 | 62.8% | 1.55 | 0.89 | 62.2% | 1.58 | 0.90 | 52.6% | 1.42 | 0.60 | 23.0% |
| Magnesium | 1.92 | 0.24 | 32.3% | 1.92 | 0.24 | 32.5% | 1.81 | 0.23 | 28.2% | 1.89 | 0.26 | 32.6% |
| MAP | 88.57 | 11.01 | 0.1% | 88.88 | 10.93 | 0.0% | 88.62 | 11.35 | 0.0% | 89.52 | 9.84 | 0.5% |
| MCH | 29.58 | 2.67 | 0.6% | 29.60 | 2.65 | 0.7% | 29.53 | 2.79 | 0.6% | 28.67 | 2.36 | 1.9% |
| MCHC | 33.14 | 1.43 | 0.6% | 33.14 | 1.41 | 0.7% | 32.56 | 1.41 | 0.6% | 32.42 | 1.33 | 1.9% |
| MCV | 89.22 | 6.57 | 0.6% | 89.27 | 6.58 | 0.7% | 90.62 | 6.98 | 0.6% | 88.43 | 6.17 | 1.9% |
| MPV | 10.21 | 0.94 | 1.7% | 10.20 | 0.94 | 1.9% | 10.24 | 0.96 | 1.4% | 10.27 | 0.92 | 2.6% |
| Oxygen supplement. | 1.97 | 36.78 | 0.0% | 1.82 | 3.14 | 0.0% | 1.84 | 3.61 | 0.0% | 3.98 | 10.28 | 0.0% |
| Phosphorus | 3.53 | 0.87 | 42.7% | 3.53 | 0.88 | 42.7% | 3.63 | 0.92 | 42.6% | 3.49 | 1.11 | 51.2% |
| Platelets | 226.13 | 101.24 | 0.7% | 227.64 | 102.49 | 0.7% | 229.58 | 104.27 | 0.6% | 229.71 | 94.47 | 1.9% |
| Potassium | 4.20 | 0.42 | 0.6% | 4.21 | 0.42 | 0.6% | 4.20 | 0.42 | 0.4% | 4.15 | 0.47 | 1.2% |
| Protein level | 6.39 | 0.88 | 34.2% | 6.37 | 0.88 | 34.1% | 6.00 | 0.82 | 30.2% | 6.03 | 0.65 | 5.3% |
| PT | 12.69 | 5.39 | 42.9% | 12.57 | 5.23 | 43.2% | 12.31 | 4.32 | 44.3% | 11.96 | 4.64 | 47.9% |
| PTT | 28.41 | 7.72 | 52.4% | 28.10 | 7.28 | 52.4% | 26.96 | 7.01 | 53.7% | 28.41 | 8.86 | 54.4% |
| Pulse rate | 80.26 | 13.03 | 0.1% | 80.56 | 12.98 | 0.0% | 80.76 | 12.78 | 0.0% | 85.82 | 13.56 | 0.2% |
| Pulse pressure | 59.28 | 12.88 | 0.1% | 59.48 | 12.84 | 0.0% | 59.67 | 12.76 | 0.0% | 57.90 | 11.34 | 0.5% |
| Race (is White) | 0.82 | 0.38 | 0.0% | 0.82 | 0.39 | 0.0% | 0.81 | 0.40 | 0.0% | 0.41 | 0.49 | 0.0% |
| Race (is Black) | 0.12 | 0.32 | 0.0% | 0.12 | 0.32 | 0.0% | 0.13 | 0.33 | 0.0% | 0.42 | 0.49 | 0.0% |
| Race (is Asian) | 0.02 | 0.14 | 0.0% | 0.02 | 0.14 | 0.0% | 0.02 | 0.14 | 0.0% | 0.06 | 0.24 | 0.0% |
| Race (is Other) | 0.04 | 0.20 | 0.0% | 0.04 | 0.20 | 0.0% | 0.05 | 0.21 | 0.0% | 0.10 | 0.31 | 0.0% |
| RBC | 3.92 | 0.74 | 0.6% | 3.92 | 0.74 | 0.7% | 3.90 | 0.77 | 0.6% | 4.24 | 0.75 | 1.9% |
| RDW | 14.70 | 2.38 | 0.7% | 14.70 | 2.39 | 0.7% | 14.77 | 2.49 | 0.6% | 14.21 | 2.07 | 1.9% |
| Resp. rate | 17.50 | 1.54 | 0.1% | 17.51 | 1.54 | 0.0% | 17.43 | 1.47 | 0.0% | 20.83 | 3.44 | 0.2% |
| Shock index | 0.65 | 0.15 | 0.1% | 0.65 | 0.14 | 0.0% | 0.65 | 0.14 | 0.0% | 0.68 | 0.13 | 0.5% |
| Shock index (age adjusted) | 36.37 | 12.39 | 0.1% | 36.48 | 12.42 | 0.0% | 36.79 | 12.33 | 0.0% | 41.72 | 11.67 | 0.5% |
| Sodium | 138.63 | 3.12 | 0.6% | 138.65 | 3.09 | 0.6% | 138.38 | 3.06 | 0.4% | 137.62 | 3.60 | 1.2% |
| SpO<sub>2</sub> | 93.81 | 3.26 | 0.1% | 93.85 | 3.20 | 0.0% | 93.67 | 3.28 | 0.0% | 91.38 | 3.65 | 0.2% |
| Systolic blood pressure | 128.09 | 17.28 | 0.1% | 128.53 | 17.17 | 0.0% | 128.40 | 17.15 | 0.0% | 128.12 | 14.73 | 0.5% |
| Temperature | 36.74 | 0.32 | 0.1% | 36.75 | 0.29 | 0.1% | 36.76 | 0.30 | 0.0% | 37.16 | 0.43 | 0.7% |
| Urine output | 309.26 | 196.66 | 12.2% | 305.09 | 188.94 | 11.9% | 287.21 | 189.28 | 15.3% | 232.61 | 170.14 | 21.9% |
| Weight | 186.13 | 54.90 | 2.1% | 186.31 | 54.69 | 2.0% | 187.38 | 56.27 | 1.4% | 199.46 | 55.81 | 3.5% |
| WBC | 9.09 | 7.60 | 0.6% | 9.32 | 9.76 | 0.7% | 9.07 | 7.05 | 0.5% | 6.90 | 3.47 | 1.9% |

## Notes

### Author Declarations

The study protocol was approved by the University of Michigan's Institutional Review Board (HUM00092309).

## References

[1] L. Wynants et al., “Prediction models for diagnosis and prognosis of covid-19 infection: systematic review and critical appraisal,” BMJ, vol. 369, p. m1328, 07 2020, doi: 10.1136/bmj.m1328.

[2] M. E. B. Smith et al., “Early warning system scores for clinical deterioration in hospitalized patients: a systematic review,” Ann. Am. Thorac. Soc., vol. 11, no. 9, pp. 1454–1465, Nov. 2014, doi: 10.1513/AnnalsATS.201403-102OC.

[3] M. D. Le Lagadec and T. Dwyer, “Scoping review: The use of early warning systems for the identification of in-hospital patients at risk of deterioration,” Aust. Crit. Care Off. J. Confed. Aust. Crit. Care Nurses, vol. 30, no. 4, pp. 211–218, Jul. 2017, doi: 10.1016/j.aucc.2016.10.003.

[4] J. McGaughey, F. Alderdice, R. Fowler, A. Kapila, A. Mayhew, and M. Moutray, “Outreach and Early Warning Systems (EWS) for the prevention of intensive care admission and death of critically ill adult patients on general hospital wards,” Cochrane Database Syst. Rev., no. 3, p. CD005529, Jul. 2007, doi: 10.1002/14651858.CD005529.pub2.

[5] “Adverse Events in Hospitals: National Incidence Among Medicare Beneficiaries - 10-Year Update,” Dec. 17, 2018. https://oig.hhs.gov/reports-and-publications/workplan/summary/wp-summary-0000328.asp (accessed Jun. 17, 2020).

[6] D. T. Linnen, G. J. Escobar, X. Hu, E. Scruth, V. Liu, and C. Stephens, “Statistical Modeling and Aggregate-Weighted Scoring Systems in Prediction of Mortality and ICU Transfer: A Systematic Review,” J. Hosp. Med., vol. 14, no. 3, pp. 161–169, 2019, doi: 10.12788/jhm.3151.

[7] S. R. Bapoje, J. L. Gaudiani, V. Narayanan, and R. K. Albert, “Unplanned transfers to a medical intensive care unit: causes and relationship to preventable errors in care,” J. Hosp. Med., vol. 6, no. 2, pp. 68–72, Feb. 2011, doi: 10.1002/jhm.812.

[8] M. P. Young, V. J. Gooder, K. McBride, B. James, and E. S. Fisher, “Inpatient transfers to the intensive care unit: delays are associated with increased mortality and morbidity,” J. Gen. Intern. Med., vol. 18, no. 2, pp. 77–83, Feb. 2003, doi: 10.1046/j.1525-1497.2003.20441.x.

[9] M. J. Rothman, “The Emperor Has No Clothes,” Crit. Care Med., vol. 47, no. 1, pp. 129–130, 2019, doi: 10.1097/CCM.0000000000003505.

[10] “Artificial Intelligence from Epic Triggers Fast, Lifesaving Care for COVID-19 Patients.” https://www.epic.com/epic/post/artificial-intelligence-epic-triggers-fast-lifesaving-care-covid-19-patients (accessed Jun. 18, 2020).

[11] K. Singh et al., “Evaluating a Widely Implemented Proprietary Deterioration Index Model Among Hospitalized COVID-19 Patients,” medRxiv, p. 2020.04.24.20079012, Jun. 2020, doi: 10.1101/2020.04.24.20079012.

[12] E. Strickland, “AI Can Help Hospitals Triage COVID-19 Patients - IEEE Spectrum,” IEEE Spectrum: Technology, Engineering, and Science News, Apr. 17, 2020. https://spectrum.ieee.org/the-human-os/artificial-intelligence/medical-ai/ai-can-help-hospitals-triage-covid19-patients (accessed Jun. 19, 2020).

[13] C. E. Gillies et al., “Demonstrating the Consequences of Learning Missingness Patterns in Early Warning Systems for Preventative Health Care: A Novel Simulation and Solution,” medRxiv, p. 2020.06.05.20123323, Jun. 2020, doi: 10.1101/2020.06.05.20123323.

[14] S. M. Lundberg et al., “From local explanations to global understanding with explainable AI for trees,” Nat. Mach. Intell., vol. 2, no. 1, Art. no. 1, Jan. 2020, doi: 10.1038/s42256-019-0138-9.

[15] M. M. Churpek et al., “Multicenter development and validation of a risk stratification tool for ward patients,” Am. J. Respir. Crit. Care Med., vol. 190, no. 6, pp. 649–655, Sep. 2014, doi: 10.1164/rccm.201406-1022OC.

[16] T. Chen and C. Guestrin, “XGBoost: A Scalable Tree Boosting System,” in Proceedings of the 22nd ACM SIGKDD International Conference on Knowledge Discovery and Data Mining, San Francisco, California, USA, Aug. 2016, pp. 785–794, doi: 10.1145/2939672.2939785.

[17] G. B. Smith, D. R. Prytherch, P. Meredith, P. E. Schmidt, and P. I. Featherstone, “The ability of the National Early Warning Score (NEWS) to discriminate patients at risk of early cardiac arrest, unanticipated intensive care unit admission, and death,” Resuscitation, vol. 84, no. 4, pp. 465–470, Apr. 2013, doi: 10.1016/j.resuscitation.2012.12.016.

[18] Royal College of Physicians, “National Early Warning Score (NEWS): Standardising the assessment of acute illness severity in the NHS.,” R. Coll. Physicians Lond. 2012.

[19] M. A. F. Pimentel et al., “A comparison of the ability of the National Early Warning Score and the National Early Warning Score 2 to identify patients at risk of in-hospital mortality: A multi-centre database study,” Resuscitation, vol. 134, pp. 147–156, Jan. 2019, doi: 10.1016/j.resuscitation.2018.09.026.

[20] R. Elshawi, M. H. Al-Mallah, and S. Sakr, “On the interpretability of machine learning-based model for predicting hypertension,” BMC Med. Inform. Decis. Mak., vol. 19, no. 1, p. 146, 29 2019, doi: 10.1186/s12911-019-0874-0.

[21] S. M. Lundberg and S.-I. Lee, “A Unified Approach to Interpreting Model Predictions,” in Advances in Neural Information Processing Systems 30, I. Guyon, U. V. Luxburg, S. Bengio, H. Wallach, R. Fergus, S. Vishwanathan, and R. Garnett, Eds. Curran Associates, Inc., 2017, pp. 4765–4774.

